# Educational Inequality in Multimorbidity: Causality and Causal Pathways. A Mendelian Randomisation Study in UK Biobank

**DOI:** 10.1101/2022.06.14.22276388

**Authors:** Teri-Louise North, Sean Harrison, Deborah C Bishop, Robyn Wootton, Alice R Carter, Tom G Richardson, Rupert A Payne, Chris Salisbury, Laura D Howe

## Abstract

**Objectives:** Multimorbidity, typically defined as having two or more long-term health conditions, is a common patient characteristic that is associated with reduced wellbeing and life expectancy. Understanding the determinants of multimorbidity may help with the design and prioritisation of interventions to prevent multimorbidity. This study examined potential causal determinants (education, BMI, smoking and alcohol consumption) of multimorbidity, and assessed the extent to which BMI, smoking and alcohol consumption explain observed educational inequalities in multimorbidity.

**Design:** Mendelian randomization study; an approach that uses genetic variants as instrumental variables to interrogate causality.

**Participants:** 181,214 females and 155,677 males, mean ages 56.7 and 57.1 years respectively, from UK Biobank.

**Main outcome measures:** Multimorbidity status (2+ self-reported health conditions); secondary analyses considered complex multimorbidity defined as 3+ or 4+ conditions, and a continuous multimorbidity score.

**Results:** Mendelian randomization suggests that lower education, higher BMI and higher levels of smoking causally increase the risk of multimorbidity. For example, one standard deviation (equivalent to 5.1 years) increase in years of education decreases the risk of multimorbidity by 9.0% (95% CI: 6.5 to 11.4%). A 5 kg/m^2^ increase in BMI is associated with a 9.2% increased risk of multimorbidity (95% CI: 8.1 to 10.3%) and a one SD higher lifetime smoking index is associated with a 6.8% increased risk of multimorbidity (95% CI: 3.3 to 10.4%). Evidence for a causal effect of alcohol consumption on multimorbidity was less strong; an increase of 5 units of alcohol per week increases the risk of multimorbidity (2+ conditions) by 1.3% (95% CI: 0.2 to 2.5%). The proportions of the association between education and multimorbidity explained by BMI and smoking are 20.4% and 17.6% respectively. Collectively, BMI and smoking account for 31.8% of the educational inequality in multimorbidity.

**Conclusions:** Education, BMI, smoking and alcohol consumption are intervenable risk factors that our results suggest have a causal effect on multimorbidity. Furthermore, BMI and lifetime smoking make a considerable contribution to the generation of educational inequalities in multimorbidity. Public health interventions that improve population-wide levels of these risk factors are likely to reduce multimorbidity and inequalities in its occurrence.

**SUMMARY BOX:** *SECTION 1: WHAT IS ALREADY KNOWN ON THIS TOPIC:*

- Multimorbidity has several known lifestyle and anthropometric risk factors and is associated with deprivation.
- The effect of education (proxying deprivation) on multimorbidity is likely mediated by some of these intervenable risk factors.
- These associations are likely to be confounded and their causality is not well understood.

*SECTION 2: WHAT THIS STUDY ADDS:*

- Analyses using genetically predicted effects suggest that education, BMI, smoking and alcohol consumption each have a causal effect on multimorbidity and that 32% of the educational inequality in multimorbidity is attributable to BMI and smoking combined.

## INTRODUCTION

Multimorbidity, defined as patients living with two or more chronic health conditions, is associated with reduced quality of life and life expectancy(1). The ageing population is driving an increase in the prevalence of multimorbidity, which already affects approximately one in four of the population in the UK and USA(2, 3). Identifying the main reversible causes of multimorbidity could inform the design of preventative strategies, helping to improve quality of life for patients and reduce the economic impact of multimorbidity.

There are profound inequalities in multimorbidity. People from more deprived backgrounds are more likely to be multimorbid, and more likely to develop multimorbidity at an earlier age. For example, a study covering one third of the Scottish population found that young and middle-aged adults in the most deprived areas often had comparable sex-specific rates of multimorbidity to those in the least deprived areas who were 10-15 years older(2). Other risk factors have been postulated for multimorbidity, including alcohol, smoking and BMI(4). Given the social patterning of these exposures, however, associations are likely to be highly confounded and establishing causality is challenging. In addition, the association of education and multimorbidity may be mediated by these risk factors.

Mendelian Randomisation (MR) is an instrumental variable (IV) analysis implemented using genetic variants robustly associated with an exposure to estimate the causal effect of the exposure on an outcome less prone to confounding and reverse causation bias(5). For an introduction to MR analysis see (6). In brief, IV analyses use another variable to proxy the exposure of interest. This ‘instrument’ is chosen because it meets strict statistical criteria and the IV estimate of the exposure-outcome association is much less likely to be biased. More recently, the MR arena has undergone rapid development(7), with new methods available to assess causality for both mediation (the causal pathways linking an exposure to an outcome)(8) and effect modification (the study of whether one exposure alters the effect of another)(9).

In this paper, we derive multimorbidity status in UK Biobank (UKBB) using self-reported data from baseline. We use MR to evaluate the causal effects of BMI, smoking, alcohol intake and years of education on multimorbidity. We evaluate the degree to which BMI, smoking, and alcohol consumption explain educational inequalities in multimorbidity, and we consider whether the risk factors interact with one another in their effects on multimorbidity.

## METHODS

### Data

UK Biobank is a population-based health research resource consisting of approximately 500,000 people, aged between 38 years and 73 years, who were recruited between the years 2006 and 2010 from across the UK(10). Particularly focused on identifying determinants of human diseases in middle-aged and older individuals, participants provided a range of information (such as demographics, health status, lifestyle measures, cognitive testing, personality self-report, and physical and mental health measures) via questionnaires and interviews; anthropometric measures, BP readings and samples of blood, urine and saliva were also taken (data available at www.ukbiobank.ac.uk). A full description of the study design, participants and quality control (QC) methods have been described in detail previously(11). UK Biobank received ethical approval from the Research Ethics Committee (REC reference for UK Biobank is 11/NW/0382).

Exposures were all assessed at the baseline research assessment. We followed a published approach (12) for inferring years of education from highest achieved qualification. Body Mass Index (BMI) in kg/m^2^ was calculated using height and weight measurements. We derived a lifetime smoking index, representing a continuous score of smoking behaviours and incorporating smoking initiation, duration, heaviness, and cessation, using a previously published approach(13, 14). (This approach was used because lifetime smoking scores incorporate heaviness but are applicable to both smokers and non-smokers.) Following the approach used in a previous Genome-Wide Association Study (GWAS)(15), we derived estimated units of alcohol consumed per week. We used responses to the baseline touchscreen questionnaire on weekly red wine, white wine and champagne, beer and cider, fortified wine, spirit and other consumption to estimate the typical units of alcohol consumed per week. Former drinkers (those who previously drank alcohol but no longer do) were set to missing and excluded from analyses, as were individuals with very high current alcohol consumption (>200 units per week). Responders who indicated they were never-drinkers were set to 0 units per week.

Our primary outcome was the standard definition of multimorbidity, the presence of two or more chronic conditions. Three additional multimorbidity measures were used in secondary analyses; the presence of 3+ and 4+ conditions, and the Cambridge multimorbidity score (CMMS) with general-outcome weights(16). This general CMMS is a continuous measure, with conditions weighted according to the average standardised weights from models of consultations, mortality and emergency admissions. For all measures of multimorbidity, the presence or absence of 35 health conditions were considered as per Payne *et al*(16) (see Condition Definitions table, Supplement). Blindness/low vision and learning disability were excluded from the original condition list owing to the lack of appropriate self-reported variables. In contrast to the condition definitions applied by Payne *et* al.(16), which included temporal restrictions and use of medications, our definitions were simplified to self-reported ‘ever’ having had a condition with the exception of cancer (self-reported doctor diagnosed new cancer estimated to be within the last 5 years, excluding non-melanoma skin cancer), hearing loss, constipation and painful condition (see supplement for full details). The information was obtained via a touchscreen questionnaire which was followed by a nurse-led interview to clarify and categorise conditions correctly. We derived each measure of multimorbidity twice, including and excluding alcohol problems in the definition, because alcohol consumption was included as an exposure or mediator in certain models. (We used the multimorbidity outcomes excluding alcohol for all models that included alcohol as an exposure/mediator.) We used the CPRD @ Cambridge – code lists (GOLD) Version 1.1 (Cambridge, UK; University of Cambridge, 2018) as a point of reference when assigning variables to condition categories, available here: https://www.phpc.cam.ac.uk/pcu/research/research-groups/crmh/cprd_cam/codelists/v11/ [downloaded May 2020].

Genetic data: Details of the in-house quality control filtering applied to the genetic data are provided in the supplement. Quality Control filtering of the UK Biobank data was conducted by R.Mitchell, G.Hemani, T.Dudding, L.Corbin, S.Harrison, L.Paternoster as described in the published protocol (doi: 10.5523/bris.1ovaau5sxunp2cv8rcy88688v)(17).

### Statistical methods

Analyses were run using Stata version 16(18) and R version 3.6.1(19). Participants were included in our analysis if they had complete data (outcome, covariates, polygenic risk score and exposure) for at least one exposure, they were of White British ancestry (to avoid confounding by population stratification) and they passed genetic QC criteria (see Supplement: Quality Control of Genetic Data). Related individuals were included in the GWAS (where relatedness was accounted for) but excluded from subsequent regression analyses. For analyses including alcohol consumption, former drinkers were removed from analyses because we were unable to consider the timing of stopping alcohol consumption in relation to the development of multimorbidity. All analyses were conducted using both standard regression models (with no instrumental variable), and using MR.

Multivariable regression analyses were used to assess the association between each exposure and each measure of multimorbidity (2+ conditions, 3+ conditions, 4+ conditions and the CMMS). Linear, rather than logistic, regression was used for all regression models so that the estimates were on the same scale as the MR estimates and represented risk differences. All multivariable regressions were run with robust standard errors and adjusted for age, sex, 40 genetic principal components and UKBB assessment centre.

MR analyses were run via two-stage least squares using *ivreg2*(20) in Stata(18) with the “robust” option specified (to enforce robust standard errors). For all MR analyses, we used a ‘split sample’ approach to avoid sample overlap with published GWASs (which can bias estimates(21)) and to implement uniform methodology across exposures. This involved splitting the UK Biobank sample into two halves randomly. Within each half, we ran a GWAS (using BOLT-LMM(22) and the MRC IEU UK Biobank GWAS pipeline https://doi.org/10.5523/bris.pnoat8cxo0u52p6ynfaekeigi) to identify genetic variants related to each of the four exposures, adjusting for age at baseline clinic, sex and 40 genetic principal components (to account for population structure). All SNPs with a p-value less than or equal to 5 × 10^−8^ were used to derive a polygenic risk score (PRS) for each exposure in the alternative split weighted by the regression coefficients from the GWAS. Clumping was performed at an R^2^ threshold of 0.001 within a 10,000 kb window, and proxies were identified using the European sub-sample of the 1000 Genomes as a reference panel(23) and a lower R^2^ limit of 0.8. The PRSs were standardised by subtracting the mean and dividing by the standard deviation. The PRSs defined based on the GWAS from one half of the UKBB sample were applied in MR analyses of the other half of the UKBB sample. MR analyses were adjusted for age, sex, 40 genetic principal components and UKBB assessment centre. The beta coefficients and standard errors from MR analyses within each half of the sample were then meta-analysed to give one estimate for beta, a 95% confidence interval, and an I^2^ estimate as an indication of heterogeneity between the estimates in each split(24, 25). Fixed-effects meta-analyses were performed using the metan command(26) in Stata. Analyses were scaled such that coefficients represented an SD change in education (equivalent to 5.1 years), a 5 kg/m^2^ increase in BMI, a 5 units per week increase in alcohol consumption, and an SD unit increase in lifetime smoking index. (As an example, a 1 SD increase in lifetime smoking is roughly the same as being a current smoker who has smoked 5 cigarettes per day for 12 years, rather than a never smoker.)

Sensitivity analyses(27) to test the assumption of no pleiotropy in MR analysis were run for the main outcome (at least two chronic conditions) (MR Egger(28), IVW(29), simple modal estimator(30) and unweighted median estimator(31)).

Mediation of the association between years of education and multimorbidity was assessed by including each potential mediator (BMI, smoking, and alcohol consumption) in turn as a covariate in a regression of multimorbidity on years of education. The joint contribution of the BMI and smoking mediators was assessed by including both variables as covariates. Similarly, in MR analyses, mediation was assessed by including both years of education and a) each mediator in turn, and b) both smoking and BMI mediators as exposures in a multivariable MR analysis(32, 33). The coefficients for years of education from these regressions and multivariable MR models estimate the ‘direct effect’; i.e. the effect of years of education on multimorbidity that operates independently of the mediator(s) being considered. The ‘indirect effect’, i.e. the effect of years of education on multimorbidity that operates through the mediator(s) is estimated by subtracting the direct effect from the total effect (the coefficient for years of education from a regression on multimorbidity not accounting for any mediators). The proportion mediated is calculated as the indirect effect divided by the total effect, multiplied by 100 to express as a percentage. 95% confidence intervals of the indirect effects and proportions mediated were calculated using Stata’s - bootstrap-command and 200 repeats. MR analyses used the same ‘split sample’ approach as the main analysis. For mediation analyses, we restricted analyses to two definitions of multimorbidity – the main outcome variable of two or more chronic conditions, and the CMMS, which, as a continuous variable, offers greatest statistical power.

Additive interaction effects between each pairwise exposure combination were assessed in multivariable linear regressions by including the product term. For MR analyses, we used a previously published approach to assessing interactions(9); for two exposures, A and B, the instruments used in the multivariable MR model are PRS exposure A, PRS exposure B, PRS exposure A x PRS exposure B, and PRS exposure A x PRS exposure A. The last instrument was included as this has been shown to be necessary in the presence of a causal effect of one exposure on the other (9). Our assumptions regarding the causal ordering of the risk factors, and hence the instruments used, are provided in the Supplement. In both multivariable regression and MR models, interactive effects were only estimated for the outcomes of multimorbidity status (2+ conditions) and the CMMS.

Stata packages used in this analysis include rsource(34), ivreg2(20) and mrrobust(27). R packages used include reshape(35), data.table(36), plyr(37), dplyr(38), R.utils(39) and devtools(40).

### Patient and Public Involvement

This study was conducted using UK Biobank. Details of patient and public involvement in the UK Biobank are available online (www.ukbiobank.ac.uk/about-biobank-uk/). No patients were specifically involved in setting the research question or the outcome measures, nor were they involved in developing plans for recruitment, design, or implementation of this study. No patients were asked to advise on interpretation or writing up of results. There are no specific plans to disseminate the results of the research to study participants or the relevant patient community, but the UK Biobank disseminates key findings from projects on its website.

## RESULTS

336,891 individuals (67% of original sample) were included in the analysis (after removal of withdrawals, those failing genetic QC/without genetic data, those without phenotype data and related individuals). In the final sample the mean age was 56.9 years (IQR 51-63 years), of whom 53.8% were female (Table 1). 55.1% of the participants had a history of at least 2 chronic conditions at baseline. 12.6% of individuals had at least 4 chronic conditions. The most common conditions overall were hearing loss (37%), anxiety & other neurotic, stress related & somatoform disorders OR depression (35%) and painful condition (29%) (Supplement page 3). The mean CMMS in the total sample was 0.7 (IQR 0.1-1.1).

**Table 1:**
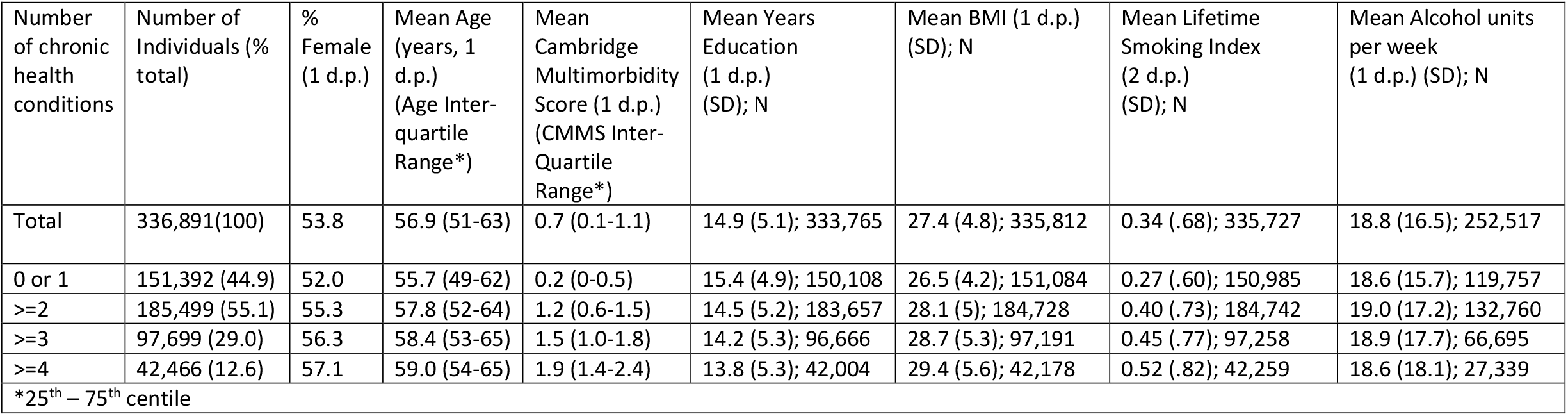
Descriptive Characteristics of Study Participants

Former drinkers (N=11,461) were removed from analyses involving alcohol. In this subset, 73% had 2+ conditions, 27% had 4+ conditions; the mean CMMS was 1.1 (1 d.p.).

### Associations of educational attainment, BMI, smoking, and alcohol consumption with multimorbidity (2+ conditions)

Both multivariable regression and MR suggest that lower years of education, higher BMI, and higher lifetime smoking index are all associated with increased risk of multimorbidity (Figure 1). In MR analyses, a one SD higher level of education (equivalent to an additional 5.1 years), is associated with a reduction in risk of multimorbidity (2+ conditions) by 9% (risk difference (RD) = -0.090, 95% CI -0.114, -0.065), a 5 kg/m^2^ increase in BMI is associated with a 9.2% increased risk of multimorbidity (RD=0.092, 95% CI=0.081, 0.103), and a one SD higher lifetime smoking index is associated with a 6.8% increased risk of multimorbidity (RD= 0.068, 95% CI=0.033, 0.104). Although both multivariable regression and MR analyses also suggest that higher alcohol consumption is a risk factor for multimorbidity, the magnitude of the effect sizes were smaller than for the other exposures. In MR analyses, an increase of 5 units of alcohol per week increases the risk of multimorbidity (2+ conditions) by 1.3% (RD=0.013, 95% CI=-0.002, 0.025). For all exposures, the estimates from MR analyses were more extreme than the estimates from multivariable regression, but the confidence intervals were wider for MR; e.g. the risk difference for multimorbidity for a 1 SD higher smoking index was 0.048 (95% CI 0.046 to 0.050) in multivariable regression, and 0.068 (95% CI 0.033 to 0.104) in MR.

**Figure 1:**
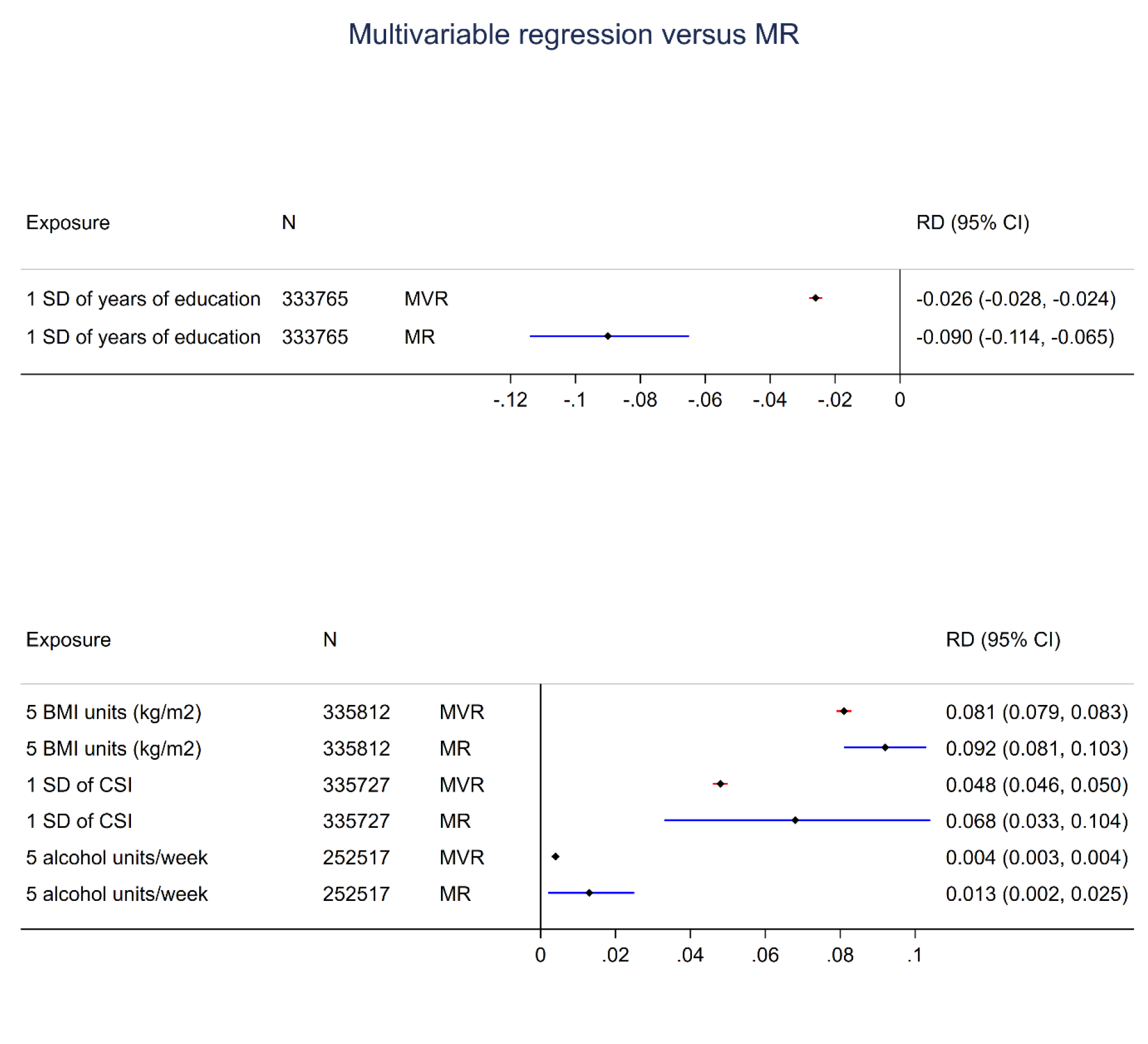
Multivariable regression (MVR) and Mendelian Randomization (MR) results for the causal effect (Risk Difference, RD) of each exposure on multimorbidity status (2+ chronic conditions)

**Figure 2:**
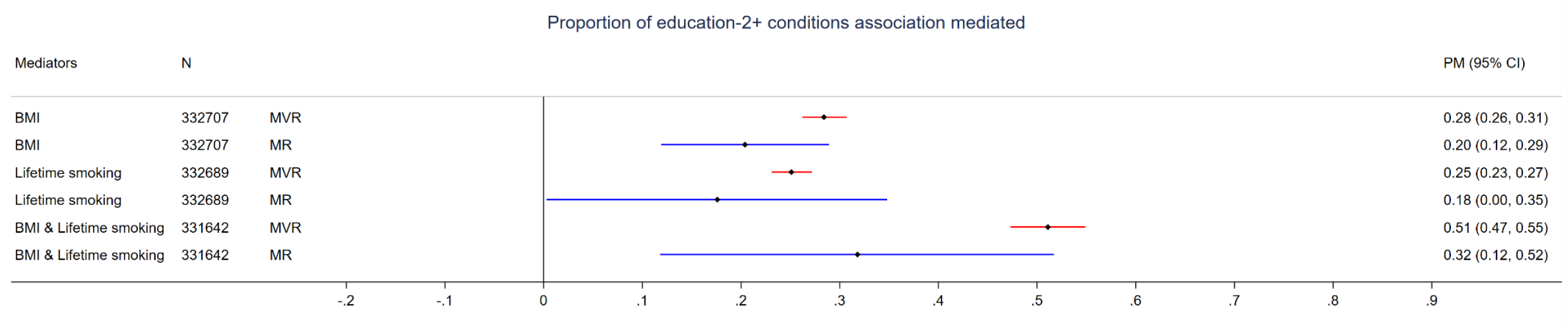
Mediation of the educational inequality in multimorbidity (2+ chronic conditions) by BMI, lifetime smoking index, and BMI and lifetime smoking index combined. Analyses conducted using multivariable regression (MVR) and Mendelian randomization (MR). Estimate presented is the Proportion Mediated (PM).

### Mechanisms explaining educational inequality in multimorbidity

In MR analyses, the proportions of the educational inequality in multimorbidity explained by BMI and smoking when each risk factor was considered separately were 20% and 18% respectively (Figure 1). When considered together in MR analyses, the two risk factors explained 32% of the educational inequality in multimorbidity. This contrasts with multivariable regression analyses, where the proportions mediated were estimated to be 28% and 25% for BMI and smoking respectively, and 51% for both risk factors combined. Multivariable regression estimated the proportion of the educational inequality in multimorbidity explained by alcohol consumption to be 0.1% (Supplementary Table 3). We did not generate an overall MR estimate for the proportion mediated by alcohol consumption because of inconsistent mediation, i.e. direct effect greater than the total effect, in one of the dataset splits (Supplementary Table 3).

### Interactions between risk factors for multimorbidity

Multivariable regression analyses to evaluate the interactive effect of pairwise combinations of the exposures on the risk of having at least 2 chronic conditions (Table 2) suggest that there are interactions between some of the risk factors, namely BMI*smoking, BMI*alcohol, smoking*alcohol, and smoking*education. However, the magnitude of all interaction terms was small. Analogous MR analyses of these interactive effects gave point estimates that were larger in magnitude than the estimates from multivariable regression, with the direction being consistent for 3/6 of the pairwise combinations, but the interactions were imprecisely estimated, with wide confidence intervals that crossed the null for all interaction terms.

**Table 2:** Interactions between risk factors for multimorbidity (2+ chronic conditions). Analyses conducted using multivariable regression (MVR) and Mendelian randomization (MR) to estimate additive interactions on the risk difference scale **

| Multivariable regression (MVR) or MR | Exposure 1 | Exposure 2 | Exposure 1 |  | Exposure 2 |  | Interaction |  |  |  |
| --- | --- | --- | --- | --- | --- | --- | --- | --- | --- | --- |
|  |  |  | Beta (3 d.p.) [I<sup>2</sup> statistic] | 95% CI (3 d.p.) | Beta (3 d.p.) [I<sup>2</sup> statistic] | 95% CI (3 d.p.) | Beta * (3 d.p.) [I<sup>2</sup> statistic] | 95% CI (3 d.p.) | p-value for interaction (3 d.p.) | N |
| MVR | 5 Units of BMI | SD Lifetime smoking index | .083 | (.082,.085) | .094 | (.085,.103) | -.009 | (-.011,-.007) | <0.001 | 334,659 |
|  | 5 Units of BMI | 5 Units of alcohol per week | .082 | (.079,.085) | .008 | (.005,.012) | -.001 | (-.002,0.000) | .002 | 251,837 |
|  | 5 Units of BMI | SD Years of education | .081 | (.076,.086) | -.015 | (-.024,-.006) | -.001 | (-.002,.001) | .460 | 332,707 |
|  | SD Lifetime smoking index | 5 Units of alcohol per week | .050 | (.047,.053) | .002 | (.001,.003) | -.001 | (-.002,-.001) | <0.001 | 251,726 |
|  | SD Lifetime smoking index | SD Years of education | .037 | (.033,.042) | -.021 | (-.023,-.019) | .003 | (.002,.005) | <0.001 | 332,689 |
|  | 5 Units of alcohol per week | SD Years of education | .004 | (.003,.006) | -.022 | (-.025,-.019) | 0.000 | (-.001,0.000) | .663 | 250,436 |
| MR | 5 Units of BMI | SD Lifetime smoking index | .175 [0.00] | (-.034,.384) | .878 [0.00] | (-1.068,2.823) | -.157 [0] | (-.520,.207) | .398 | 334,659 |
|  | 5 Units of BMI | 5 Units of alcohol per week | .074 [2.42] | (-.123,.272) | -.009 [27.33] | (-.333,.314) | .003 [24.81] | (-.053,.060) | .912 | 251,837 |
|  | 5 Units of BMI | SD Years of education | -.201 [0.00] | (-.550,.148) | -.602 [0.00] | (-1.267,.062) | .098 [0.00] | (-.023,.219) | .113 | 332,707 |
|  | SD Lifetime smoking index | 5 Units of alcohol per week | .302 [0] | (-.134,.738) | .049 [0.00] | (-.022,.119) | -.052 [0.00] | (-.150,.046) | .299 | 251,726 |
|  | SD Lifetime smoking index | SD Years of education | .131 [1.48] | (-.423,.684) | -.046 [37.99] | (-.151,.059) | -.028 [3.26] | (-.234,.178) | .789 | 332,689 |
|  | 5 Units of alcohol per week | SD Years of education | -.142 [0.00] | (-.409,.125) | -.292 [11.96] | (-.623,.040) | .052 [0.00] | (-.036,.140) | .249 | 250,436 |
| *Risk difference for interaction coefficient from linear regression, adjusted for sex, age, centre, PCs, exposure 1 and exposure 2 (MVR) or Meta-analysed estimate of interaction coefficient from two two-stage least squares estimates from split-sample analyses, adjusted for age, sex, centre and PCs (MR)<br>**Alcohol excluded from multimorbidity definition when it is part of the interaction |  |  |  |  |  |  |  |  |  |  |

### Sensitivity analyses

Secondary analyses using alternative definitions of multimorbidity yielded a similar pattern of results for the associations of years of education, BMI, smoking, and alcohol consumption with multimorbidity (Supplementary Tables 1, 2) and for mediation of the educational inequality in the CMMS (Supplementary Table 3). Similar to the main outcome, MR confidence intervals for all interaction terms were wide when analyses were repeated with CMMS as the outcome and the direction of effect was consistent with multivariate regression for 2/6 pairwise combinations (Supplementary Table 4). In multivariate regression analyses, the direction of the interactive effect of smoking and education on the CMMS was in the opposite direction compared with the main outcome.

Sensitivity analyses to test the assumption (in the main analysis) of no pleiotropy revealed estimates that were generally directionally consistent but of smaller magnitude for education and BMI. The MR-egger constant estimates suggest evidence for directional pleiotropy for BMI and smoking. The MR-Egger slope estimate was in the opposite direction to the main analysis for smoking, while the other estimates (IVW, simple modal and unweighted median) were directionally consistent but of larger magnitude.

## DISCUSSION

This study has provided evidence for a causal effect of lower educational attainment, higher BMI and higher level of smoking on multimorbidity status. There was also weak evidence for a causal effect of greater alcohol consumption on risk of multimorbidity, although the magnitude of effects was generally smaller than for the other risk factors. In our analyses, one standard deviation of years of education (equivalent to 5.1 additional years) equates approximately to a 9% decrease in risk of multimorbidity. For education, BMI, smoking, and alcohol consumption, estimated effects on multimorbidity were greater in MR analyses compared with multivariable regression. However, confidence intervals for MR results were wide and, with the exception of the coefficient for education, spanned the point estimate from multivariable regression models.

Our analysis suggests that 20% of educational inequality in multimorbidity is explained by BMI, and 32% is jointly explained by BMI and smoking. This is slightly less than the 51% of educational inequality in multimorbidity explained by BMI and smoking in multivariable regression. However, 48-88% of the total effect of education on multimorbidity remains unaccounted for by these risk factors. We did not include alcohol in conjunction with the other potential mediators because neither multivariable nor MR analyses provided evidence that alcohol consumption mediated the effect of education on multimorbidity. Units consumed per week is also a crude measure of alcohol consumption, which could partially explain the lack of mediation by alcohol use. Looking ahead we need to consider other explanatory mechanisms, which are likely to be numerous, complex and span multiple social, behavioural and biological domains.

While there may be interactions between various lifestyle and anthropometric exposures on risk of multimorbidity, we could not provide evidence for these within a causal framework possibly due to low power to detect interactive effects. In multivariable analyses, where statistical power is greater than MR, interactions were generally of small magnitude, and were most often in the opposite direction to the main effects of the risk factors (i.e. the cumulative effect of having both risk factors was generally less than would be predicted from their individual effects), suggesting that interactions between the risk factors we studied are not a major contributor to the aetiology of multimorbidity.

A recent study(41) of over 400,000 GP-registered adults in England concluded that over half of health service utilisation is attributable to individuals with multimorbidity. Furthermore, the ageing population is leading to an increase in the prevalence of multimorbidity over time. Identifying the preventable causal determinants of multimorbidity is thus paramount for easing the pressure on health services. Our analysis suggests that population-level interventions to reduce BMI and smoking would likely lead to both a reduction in the occurrence of multimorbidity, and a reduction in educational inequalities in multimorbidity.

A key strength of our study is the use of Mendelian randomization to improve causal inference. In traditional epidemiological study designs, confounding factors and reverse causation can bias the estimated associations between putative risk factors for multimorbidity. Furthermore, when analysing the mediating pathways explaining educational inequalities in multimorbidity, measurement error in the mediator can lead to an underestimate of the contribution of mediating variables(42). The use of MR overcomes these limitations of previous analyses.

There is a body of work devoted to defining multimorbidity(2, 16, 41). We explored three definitions of multimorbidity increasing in severity from 2+ to 4+ chronic conditions, in addition to a multimorbidity score, which captures all available information as a continuum with conditions weighted by the average standardised weights from models of consultations, mortality and emergency admissions. Findings were generally consistent across these definitions. Nonetheless, our findings may be driven by the prevalence of conditions feeding into the definition of multimorbidity. Multimorbidity is not a single ‘entity’; for different patients the state of multimorbidity can represent diverse combinations of health conditions. The most commonly reported health conditions in this study were hearing loss (37%), anxiety & other neurotic, stress related & somatoform disorders OR depression (35%) and painful condition (29%). Thus, our findings may represent established causal effects of education, BMI(43), smoking, and alcohol on these conditions. Nonetheless, as these conditions underlie many cases of multimorbidity, this does not detract from the implications of our findings about the potential public health and clinical impact of interventions to improve population levels of these risk factors. Further work identifying distinct clusters of health conditions to explore whether different ‘types’ of multimorbidity have distinct aetiologies may be of interest.

Our study has several limitations. Firstly, the use of self-reported health conditions may have led to misclassification of multimorbidity status for some people. However, self-reported data (unlike linked primary and secondary care data) was available across the whole sample. Secondly, UK Biobank participants are known to be over-selected from higher socio-economic categories(44), and the use of genetic data in this analysis necessitated restricting to people of White British ethnicity. This may have led to underestimation of the effects of exposures on multimorbidity, such that the effects we demonstrate can be viewed as minimal likely causal effects in a population more representative of the UK as a whole. Thirdly, with the exception of cancer, hearing loss, painful condition and constipation, we defined chronic conditions based on self-reported ever having been diagnosed by a doctor. In contrast, some studies(41) base their definition on long-term “currently active” conditions, making our definition less specific. However, our definition ensures that we can be as inclusive as possible with regards to the conditions contributing to multimorbidity. Fourthly, we excluded former drinkers from analyses of alcohol because these individuals are known to have worse health outcomes than never drinkers and analysing them as non-drinkers would be inappropriate. The available data in UK Biobank does not permit detailed analysis of prior drinking patterns or time-since stopping alcohol consumption. However, this means that our conclusions about the effects of alcohol on multimorbidity may not extend to former drinkers. In addition, removal of former drinkers reduced the sample size for these analyses and hence the power to detect effects. Although we found weak evidence of an effect of alcohol on multimorbidity, the effect size was smaller than for the other risk factors. This may at least partially reflect the complexity of defining alcohol use. Here we used a continuous measure of units per week, but other measures such as binge drinking may also be relevant for disease outcomes, particularly in the context of educational attainment(45). Our analysis of current alcohol units consumed per week also does not capture previously heavy but now light drinkers. An additional study limitation is that our analyses assume linear effects of the risk factors on multimorbidity; this assumption may not hold for all relationships. Importantly, although we checked where possible that our analysis met the assumptions(7) of MR, our conclusions rely on the validity of these assumptions. Lastly, although multivariable regression analyses demonstrated some interactions between risk factors, these were not detected in MR analyses. This likely reflects insufficient power to examine interactive effects within a causal framework.

This results of this study suggest that education, BMI, smoking, and, to a lesser degree, alcohol consumption, all have causal effects on multimorbidity. Furthermore, BMI and smoking explain approximately one third of the educational inequality in multimorbidity. In the UK, school attendance is compulsory until age 18, and policies to increase educational attendance would therefore focus on increasing university participation. Such policies may potentially influence multimorbidity risk. However, policies to mitigate the health disadvantage of low education may be more realistic and within reach of public health, thus motivating our study of the pathways explaining educational inequality in multimorbidity. Interventions to reduce population levels of BMI and smoking could lead to reduced occurrence and reduced educational inequalities in multimorbidity.

## Data Availability

All data are available on request from UK Biobank. Statistical code are currently available on request; they will shortly be published on a code repository

https://www.ukbiobank.ac.uk/enable-your-research/apply-for-access

## ACKNOWLEDGEMENTS

This research has been conducted using the UK Biobank Resource under Application Number 19278.

This work was carried out using the computational facilities of the Advanced Computing Research Centre, University of Bristol – http://www.bristol.ac.uk/acrc/.

This study used the MRC IEU UK Biobank GWAS pipeline. Please see: Elsworth, BL, Mitchell, R, Raistrick, CA, Paternoster, L, Hemani, G, Gaunt, TR (2019): MRC IEU UK Biobank GWAS pipeline version 2. https://doi.org/10.5523/bris.pnoat8cxo0u52p6ynfaekeigi

