## Supplementary materials for "Educational Inequality in Multimorbidity: Causality and Causal Pathways. A Mendelian Randomisation Study in UK Biobank"

#### Condition definitions

| | Condition name (following Payne *et al.*(1), but some differences) | Definition in this study [biobank variable code] | % |
|---|---|---|---|
| 1 | Alcohol problems | 1. Self-reported previous doctor diagnosed alcohol dependency [field 20002, baseline] | .14 |
| 2 | Anorexia or bulimia | 1. Self-reported previous doctor diagnosed anorexia/bulimia/other eating disorder [field 20002, baseline] | .07 |
| 3 | Atrial fibrillation | 2. Self-reported previous doctor diagnosed atrial fibrillation [field 20002, baseline]<br>3. Self-reported previous doctor diagnosed atrial flutter [field 20002, baseline] | .80 |
| 4 | Blindness and low vision | EXCLUDED | |
| 5 | Bronchiectasis | 1. Self-reported previous doctor diagnosed bronchiectasis [field 20002, baseline] | .23 |
| 6 | Chronic liver disease and viral hepatitis | 1. Self-reported previous doctor diagnosed hepatitis [field 20002, baseline]<br>2. Self-reported previous doctor diagnosed infective/viral hepatitis [field 20002, baseline]<br>3. Self-reported previous doctor diagnosed non-infective hepatitis [field 20002, baseline]<br>4. Self-reported previous doctor diagnosed liver failure/cirrhosis [field 20002, baseline]<br>5. Self-reported previous doctor diagnosed hepatitis b [field 20002, baseline]<br>6. Self-reported previous doctor diagnosed hepatitis c [field 20002, baseline]<br>7. Self-reported previous doctor diagnosed alcoholic liver disease/alcoholic cirrhosis [field 20002, baseline]<br>8. Self-reported previous doctor diagnosed primary biliary cirrhosis [field 20002, baseline] | .53 |
| 7 | Chronic sinusitis | 1. Self-reported previous doctor diagnosed chronic sinusitis [field 20002, baseline]<br>2. Self-reported previous doctor diagnosed nasal/sinus disorder [field 20002, baseline]<br>3. Self-reported previous doctor diagnosed nasal polyps [field 20002, baseline] | 1.42 |
| 8 | COPD | 1. Self-reported previous doctor diagnosed chronic obstructive airways disease/copd [ field 20002, baseline]<br>2. Self-reported previous doctor diagnosed emphysema/chronic bronchitis [ field 20002, baseline]<br>3. Self-reported previous doctor diagnosed emphysema [ field 20002, baseline] | 1.82 |

| | | | |
|---|---|---|---|
| | | 4. "Has a doctor ever told you that you have had any of the following conditions?"…"emphysema/chronic bronchitis" [field 6152, baseline] | |
| 9 | Coronary heart disease | 1. Self-reported previous doctor diagnosed angina [field 20002, baseline]<br>2. Self-reported previous doctor diagnosed heart attack/myocardial infarction [field 20002, baseline]<br>3. "Has a doctor ever told you that you have had any of the following conditions?"…"Heart attack" [Field 6150, baseline]<br>4. "Has a doctor ever told you that you have had any of the following conditions?"…"Angina" [Field 6150, baseline]<br>5. Self-reported previous doctor diagnosed cardiomyopathy [field 20002, baseline]<br>6. Self-reported previous doctor diagnosed hypertrophic cardiomyopathy [field 20002, baseline]<br>7. Self-reported previous doctor diagnosed aortic aneurysm [field 20002, baseline]<br>8. Self-reported previous doctor diagnosed aortic aneurysm rupture [field 20002, baseline]<br>9. Self-reported previous doctor diagnosed aortic valve disease [field 20002, baseline]<br>10. Self-reported previous doctor diagnosed aortic regurgitation/incompetence [field 20002, baseline] | 4.65 |
| 10 | Dementia or cognitive impairment | 1. Self-reported previous doctor diagnosed dementia/alzheimers/cognitive impairment [field 20002, baseline] | .02 |
| 11 | Diabetes | 1. Self-reported previous doctor diagnosed diabetic eye disease [field 20002, baseline]<br>2. "Has a doctor ever told you that you have diabetes?" [field 2443, baseline]<br>3. Self-reported previous doctor diagnosed diabetes [field 20002, baseline]<br>4. Self-reported previous doctor diagnosed type I diabetes [field 20002, baseline]<br>5. Self-reported previous doctor diagnosed type II diabetes [field 20002, baseline]<br>6. Self-reported previous doctor diagnosed diabetic neuropathy/ulcers [field 20002, baseline]<br>7. Self-reported previous doctor diagnosed diabetic nephropathy [field 20002, baseline] | 4.81 |
| 12 | Diverticular disease of intestine | 1. Self-reported previous doctor diagnosed diverticular disease/diverticulitis [field 20002, baseline] | 1.13 |
| 13 | Hearing loss | 1. "Do you have any difficulty with your hearing?"..."I am completely deaf" [field 2247, baseline]<br>2. "Do you use a hearing aid most of the time?" [field 3393, baseline] | 37.28 |

| | | | |
|---|---|---|---|
| | | 3. "Do you find it difficult to follow a conversation if there is background noise (such as TV, radio, children playing?)" [field 2257, baseline]<br>4. Self-reported previous doctor diagnosed otosclerosis [field 20002, baseline]<br>5. Self-reported previous doctor diagnosed meniere's disease [field 20002, baseline] | |
| 14 | Heart failure | 1. Self-reported previous doctor diagnosed heart failure/pulmonary oedema [field 20002, baseline] | .07 |
| 15 | Hypertension | 1. Self-reported previous doctor diagnosed hypertension [field 20002, baseline]<br>2. Self-reported previous doctor diagnosed essential hypertension [field 20002, baseline]<br>3. "Has a doctor ever told you that you have had any of the following conditions?" … "High blood pressure" [field 6150 baseline] | 27.39 |
| 16 | Inflammatory bowel disease | 1. Self-reported previous doctor diagnosed inflammatory bowel disease [field 20002, baseline]<br>2. Self-reported previous doctor diagnosed crohns disease [field 20002, baseline]<br>3. Self-reported previous doctor diagnosed ulcerative colitis [field 20002, baseline]<br>4. Self-reported previous doctor diagnosed colitis/ not crohns or ulcerative colitis [field 20002, baseline] | 1.12 |
| 17 | Learning disability | EXCLUDED | |
| 18 | Multiple sclerosis | 1. Self-reported previous doctor diagnosed multiple sclerosis [field 20002, baseline] | .37 |
| 19 | Parkinson's disease | 1. Self-reported previous doctor diagnosed parkinson's disease [field 20002, baseline] | .18 |
| 20 | Peptic ulcer disease | 1. Self-reported previous doctor diagnosed peptic ulcer [field 20002, baseline]<br>2. Self-reported previous doctor diagnosed duodenal ulcer [field 20002, baseline]<br>3. Self-reported previous doctor diagnosed gastric/stomach ulcers [field 20002, baseline] | 1.15 |
| 21 | Peripheral vascular disease | 1. Self-reported previous doctor diagnosed peripheral vascular disease [field 20002, baseline]<br>2. Self-reported previous doctor diagnosed leg claudication/intermittent claudication [field 20002, baseline] | .26 |
| 22 | Prostate disorders | 1. Self-reported previous doctor diagnosed prostate problem (not cancer) [field 20002, baseline]<br>2. Self-reported previous doctor diagnosed enlarged prostate [field 20002, baseline]<br>3. Self-reported previous doctor diagnosed benign prostatic hypertrophy [field 20002, baseline]<br>4. Self-reported previous doctor diagnosed prostatitis [field 20002, baseline] | 1.73 |
| 23 | Psychoactive substance misuse (not alcohol) | 1. Self-reported previous doctor diagnosed opioid dependency [field 20002, baseline] | .02 |

| | | | |
|---|---|---|---|
| | | 2. Self-reported previous doctor diagnosed other substance abuse/dependency [field 20002, baseline] | |
| 24 | Rheumatoid arthritis, other inflammatory polyarthropathies & systemic connective tissue disorders | 1. Self-reported previous doctor diagnosed connective tissue disorder [field 20002, baseline]<br>2. Self-reported previous doctor diagnosed rheumatoid arthritis [field 20002, baseline]<br>3. Self-reported previous doctor diagnosed systemic lupus erythematosis [field 20002, baseline]<br>4. Self-reported previous doctor diagnosed psoriatic arthropathy [field 20002, baseline]<br>5. Self-reported previous doctor diagnosed ankylosing spondylitis [field 20002, baseline]<br>6. Self-reported previous doctor diagnosed polymyalgia rheumatica [field 20002, baseline]<br>7. Self-reported previous doctor diagnosed myositis/myopathy [field 20002, baseline]<br>8. Self-reported previous doctor diagnosed dermatopolymyositis [field 20002, baseline]<br>9. Self-reported previous doctor diagnosed dermatomyositis [field 20002, baseline]<br>10. Self-reported previous doctor diagnosed polymyositis [field 20002, baseline]<br>11. Self-reported previous doctor diagnosed scleroderma/ systemic sclerosis [field 20002, baseline]<br>12. Self-reported previous doctor diagnosed vasculitis [field 20002, baseline] | 2.02 |
| 25 | Stroke and TIA | 1. Self-reported previous doctor diagnosed stroke [field 20002, baseline]<br>2. Self-reported previous doctor diagnosed transient ischaemic attack [field 20002, baseline]<br>3. Self-reported previous doctor diagnosed ischaemic stroke [field 20002, baseline]<br>4. “Has a doctor ever told you that you have had any of the following conditions?” … “Stroke” [field 6150 baseline]<br>5. Self-reported previous doctor diagnosed brain haemorrhage [field 20002, baseline] | 1.71 |
| 26 | Thyroid disorders | 1. Self-reported previous doctor diagnosed hypothyroidism/myxoedema [field 20002, baseline]<br>2. Self-reported previous doctor diagnosed hyperthyroidism/thyrotoxicosis [field 20002, baseline]<br>3. Self-reported previous doctor diagnosed thyroid goitre [field 20002, baseline]<br>4. Self-reported previous doctor diagnosed grave’s disease [field 20002, baseline] | 5.50 |
| 27 | Constipation | 1. Self-reported previous doctor diagnosed constipation [field 20002, baseline] | 2.85 |

| | | | |
|---|---|---|---|
| | | 2. "Do you regularly take any of the following"…."Laxatives (e.g. Dulcolax, Senokot)" [field 6154, baseline] | |
| 28 | Migraine | 1. Self-reported previous doctor diagnosed migraine [field 20002, baseline] | 2.96 |
| 29 | Epilepsy | 1. Self-reported previous doctor diagnosed epilepsy [field 20002, baseline] | .84 |
| 30 | Asthma | 1. Self-reported previous doctor diagnosed asthma [field 20002, baseline]<br>2. "Has a doctor ever told you that you have had any of the following conditions?"…"Asthma" [field 6152, baseline] | 11.70 |
| 31 | Irritable bowel syndrome | 1. Self-reported previous doctor diagnosed irritable bowel syndrome [field 20002, baseline] | 2.37 |
| 32 | Psoriasis or Eczema | 1. Self-reported previous doctor diagnosed contact dermatitis [field 20002, baseline]<br>2. Self-reported previous doctor diagnosed eczema/dermatitis [field 20002, baseline]<br>3. Self-reported previous doctor diagnosed psoriasis [field 20002, baseline] | 3.66 |
| 33 | Anxiety and other neurotic, stress related and somatoform disorders OR depression | 1. "Have you ever seen a psychiatrist for nerves, anxiety, tension or depression?" [field 2100, baseline]<br>2. "Have you ever seen a general practitioner (GP) for nerves, anxiety, tension or depression?" [field 2090, baseline]<br>3. Self-reported previous doctor diagnosed post-traumatic stress disorder [field 20002, baseline]<br>4. Self-reported previous doctor diagnosed obsessive compulsive disorder (ocd) [field 20002, baseline]<br>5. Self-reported previous doctor diagnosed depression [field 20002, baseline]<br>6. Self-reported previous doctor diagnosed anxiety/panic attacks [field 20002, baseline]<br>Self-reported previous doctor diagnosed nervous breakdown [field 20002, baseline] | 34.98 |
| 34 | Cancer – [New] diagnosis in last five years (excluding non-melanoma skin cancer) | Self -reported doctor diagnosed cancer, estimated to have first occurred in the last 5 years, excluding non-melanoma skin cancer [field 20007, baseline; field 20001, baseline] | 3.62 |
| 35 | Chronic kidney disease | 1. Self-reported previous doctor diagnosed renal/kidney failure [field 20002, baseline]<br>2. Self-reported previous doctor diagnosed renal failure requiring dialysis [field 20002, baseline]<br>3. Self-reported previous doctor diagnosed renal failure not requiring dialysis [field 20002, baseline]<br>4. Self-reported previous doctor diagnosed kidney nephropathy [field 20002, baseline]<br>5. Self-reported previous doctor diagnosed diabetic nephropathy [field 20002, baseline] | .18 |

| | | | |
|---|---|---|---|
| 36 | Painful condition | 1. "Do you regularly take any of the following"…."Ibuprofen (e.g. Nurofen)" [field 6154, baseline]<br>2. "Do you regularly take any of the following"…."Paracetamol" [field 6154, baseline] | 29.48 |
| 37 | Schizophrenia (and related non-organic psychosis) or bipolar disorder | 1. Self-reported previous doctor diagnosed schizophrenia [field 20002, baseline]<br>2. Self-reported previous doctor diagnosed mania/ bipolar disorder/ manic depression [field 20002, baseline] | .36 |

As a reference we used the CPRD @ Cambridge – code lists (GOLD) Version 1.1 when assigning variables to condition categories, available here https://www.phpc.cam.ac.uk/pcu/research/research-groups/crmh/cprd_cam/codelists/v11/

[downloaded May 2020]

#### Quality Control of Genetic Data

Quality Control filtering of the UK Biobank data was conducted by R.Mitchell, G.Hemani, T.Dudding, L.Corbin, S.Harrison, L.Paternoster as described in the published protocol (doi: 10.5523/bris.1ovaau5sxunp2cv8rcy88688v)(2).

The full data release contains the cohort of successfully genotyped samples (n=488,377). 49,979 individuals were genotyped using the UK BiLEVE array and 438,398 using the UK Biobank axiom array. Pre-imputation QC, phasing and imputation are described elsewhere(3). In brief, prior to phasing, multiallelic SNPs or those with MAF ≤1% were removed. Phasing of genotype data was performed using a modified version of the SHAPEIT2 algorithm(4). Genotype imputation to a reference set combining the UK10K haplotype and HRC reference panels(5) was performed using IMPUTE2 algorithms(6). The analyses presented here were restricted to autosomal variants using a graded filtering with varying imputation quality for different allele frequency ranges. Therefore, rarer genetic variants are required to have a higher imputation INFO score (Info>0.3 for MAF >3%; Info>0.6 for MAF 1-3%; Info>0.8 for MAF 0.5-1%; Info>0.9 for MAF 0.1-0.5%) with MAF and Info scores having been recalculated on an in-house derived 'European' subset. Data quality control Individuals with sex-mismatch (derived by comparing genetic sex and reported sex) or individuals with sex-chromosome aneuploidy were excluded from the analysis. Ancestry: We restricted the sample to individuals of white British ancestry who self-report as "White British" and who have very similar ancestral backgrounds according to the PCA, as described by Bycroft(3).

Degree of relatedness: Estimated kinship coefficients using the KING toolset(7) identified 107,162 pairs of related individuals(3). We did not remove related study members from our analysis with the exception of 2 individuals who were related to a very large number (>200) of individuals.

#### Assumptions of the causal ordering of risk factors

Smoking -> BMI -> Multimorbidity

BMI -> Alcohol -> Multimorbidity

Education -> BMI -> Multimorbidity

Alcohol -> Smoking -> Multimorbidity

Education -> Smoking -> Multimorbidity

Education -> Alcohol -> Multimorbidity

#### Results Tables

Table 1: Multivariable regression risk differences from associations of exposures with Multimorbidity Status (2+, 3+, 4+ conditions) and CMMS

| Exposure | Outcome | Beta* (3 d.p.) | 95% CI (3 d.p.) | p-value (3 d.p.) | N |
|---|---|---|---|---|---|
| SD Years of education | 2+ conditions | -.026 | (-.028,-.024) | <0.001 | 333,765 |
| | 3+ conditions | -.026 | (-.028,-.025) | <0.001 | 333,765 |
| | 4+ conditions | -.018 | (-.020,-.017) | <0.001 | 333,765 |
| | CMMS | -.059 | (-.062,-.056) | <0.001 | 333,765 |
| 5 Units of BMI | 2+ conditions | .081 | (.079,.083) | <0.001 | 335,812 |
| | 3+ conditions | .081 | (.079,.082) | <0.001 | 335,812 |
| | 4+ conditions | .054 | (.053,.056) | <0.001 | 335,812 |
| | CMMS | .115 | (.112,.118) | <0.001 | 335,812 |
| SD Lifetime smoking index | 2+ conditions | .048 | (.046,.050) | <0.001 | 335,727 |
| | 3+ conditions | .049 | (.047,.051) | <0.001 | 335,727 |
| | 4+ conditions | .034 | (.032,.035) | <0.001 | 335,727 |
| | CMMS | .108 | (.105,.111) | <0.001 | 335,727 |
| 5 Units of alcohol per week** | 2+ conditions | .004 | (.003,.004) | <0.001 | 252,517 |
| | 3+ conditions | .002 | (.002,.003) | <0.001 | 252,517 |
| | 4+ conditions | 0.000 | (0.000,.001) | .037 | 252,517 |
| | CMMS | .002 | (.001,.003) | <0.001 | 252,517 |

*Risk difference from linear regression, adjusted for sex, age, centre and PCs
**Alcohol excluded from multimorbidity definition when it is an exposure

#### Table 2: MR risk differences from associations of exposures with Multimorbidity Status (2+, 3+, 4+ conditions) and CMMS

| Exposure | Outcome | Beta* (3 d.p.) [$I^2$ statistic] | 95% CI (3 d.p.) | p-value (3 d.p.) | N |
|---|---|---|---|---|---|
| SD Years of education | 2+ conditions | -.090 [55.75] | (-.114,-.065) | <0.001 | 333,765 |
| | 3+ conditions | -.086 [0.00] | (-.108,-.063) | <0.001 | 333,765 |
| | 4+ conditions | -.055 [0.00] | (-.071,-.038) | <0.001 | 333,765 |
| | CMMS | -.180 [54.59] | (-.217,-.143) | <0.001 | 333,765 |
| 5 Units of BMI | 2+ conditions | .092 [44.30] | (.081,.103) | <0.001 | 335,812 |
| | 3+ conditions | .093 [75.83] | (.083,.103) | <0.001 | 335,812 |
| | 4+ conditions | .060 [0.00] | (.052,.067) | <0.001 | 335,812 |
| | CMMS | .138 [60.72] | (.122,.154) | <0.001 | 335,812 |
| SD Lifetime smoking index | 2+ conditions | .068 [0.00] | (.033,.104) | <0.001 | 335,727 |
| | 3+ conditions | .083 [0.00] | (.051,.115) | <0.001 | 335,727 |
| | 4+ conditions | .044 [62.76] | (.020,.067) | <0.001 | 335,727 |
| | CMMS | .177 [0.00] | (.125,.229) | <0.001 | 335,727 |
| 5 Units of alcohol per week** | 2+ conditions | .013 [0.00] | (.002,.025) | .022 | 252,517 |
| | 3+ conditions | .011 [0.00] | (.001,.021) | .037 | 252,517 |
| | 4+ conditions | .007 [0.00] | (0.000,.014) | .064 | 252,517 |
| | CMMS | .008 [0.00] | (-.008,.025) | .308 | 252,517 |

*Meta-analysed estimate from two two-stage least squares estimates from split-sample analyses, adjusted for age, sex, centre and PCs
**Alcohol excluded from multimorbidity definition when it is an exposure

#### Table 3: Education and multimorbidity: Proportion mediated and Indirect effects

| Exposure* | Mediators* | Outcome | Multivariate regression or MR | Proportion mediated** (95% CI) | Indirect effect** (95% CI) |
|---|---|---|---|---|---|
| Years of education | BMI | 2+ conditions | Multivariate regression | .284 (.262,.307) | -.007 (-.008,-.007) |
| | Lifetime smoking index | | | .251 (.231,.272) | -.006 (-.007,-.006) |
| | Units of alcohol per week | | | .001 (-.002,.003) | 0.000(0.000,0.000) |
| | BMI & smoking | | | .511 (.473,.549) | -.013 (-.014,-.013) |
| | BMI | | MR | .204 [39.42] (.119,.289) | -.026 [69.77] (-.031,-.02) |
| | Lifetime smoking index | | | .176 [0] (.003,.348) | -.018 [0] (-.032,-.004) |
| | Units of alcohol per week | | | -0.006(-0.074,0.063)[split 1]; -0.002(-0.039,0.035)[split 2] | 0.000 (-0.003,0.004)[split 1]; 0.000 (-0.004,0.004)[ [split 2] |
| | BMI & smoking | | | .318 [0] (.118,.517) | -.034 [0] (-.049,-.019) |
| | BMI | CMMS | Multivariate regression | .174 (.163,.184) | -.01 (-.011,-.01) |
| | Lifetime smoking index | | | .247 (.233,.26) | -.015 (-.015,-.014) |
| | Units of alcohol per week | | | 0.000(-.001,.001) | 0.000(0.000,0.000) |
| | BMI & smoking | | | .403 (.383,.422) | -.023 (-.024,-.023) |
| | BMI | | MR | .166 [74.45] (.109,.222) | -.036 [76.45] (-.044,-.029) |
| | Lifetime smoking index | | | .267 [0] (.123,.411) | -.051 [0] (-.073,-.028) |
| | Units of alcohol per week | | | -0.001(-0.026,0.024)[split 1]; -0.001(-0.024,0.023)[split 2] | 0.000 (-0.004,0.004)[split 1]; 0.000 (-0.003,0.004) [split 2] |
| | BMI & smoking | | | .358 [0] (.214,.502) | -.071 [0] (-.093,-.049) |

*All exposures and mediators scaled as described in main paper
**MR estimates have been meta-analysed across splits and include an I2 value in square brackets with the exception of mediator=alcohol, where the estimates in each split are reported due to inconsistent mediation

#### Table 4: Multivariate regression and MR risk differences from associations of interaction terms with CMMS** [Additive scale]

| Multivariate regression or MR | Exposure 1 | Exposure 2 | Exposure 1 | | Exposure 2 | | Interaction | | | |
|---|---|---|---|---|---|---|---|---|---|---|
| | | | Beta (3 d.p.) [$I^2$ statistic] | 95% CI (3 d.p.) | Beta (3 d.p.) [$I^2$ statistic] | 95% CI (3 d.p.) | Beta * (3 d.p.) [$I^2$ statistic] | 95% CI (3 d.p.) | p-value for interaction (3 d.p.) | N |
| Multivariate regression | 5 Units of BMI | SD Lifetime smoking index | .111 | (.108,.115) | .130 | (.112,.148) | -.005 | (-.008,-.002) | .002 | 334,659 |
| | 5 Units of BMI | 5 Units of alcohol per week | .113 | (.108,.118) | .021 | (.015,.027) | -.003 | (-.005,-.002) | <0.001 | 251,837 |
| | 5 Units of BMI | SD Years of education | .139 | (.130,.148) | .008 | (-.008,.025) | -.010 | (-.013,-.007) | <0.001 | 332,707 |
| | SD Lifetime smoking index | 5 Units of alcohol per week | .102 | (.097,.107) | -.002 | (-.003,-.001) | -.001 | (-.002,-.001) | <0.001 | 251,726 |
| | SD Lifetime smoking index | SD Years of education | .117 | (.109,.126) | -.041 | (-.044,-.038) | -.006 | (-.009,-.003) | <0.001 | 332,689 |
| | 5 Units of alcohol per week | SD Years of education | .002 | (-.001,.005) | -.049 | (-.053,-.044) | 0.000 | (-.001,.001) | .992 | 250,436 |
| MR | 5 Units of BMI | SD Lifetime smoking index | .228 [0] | (-.081,.537) | 1.247 [0] | (-1.622,4.117) | -.206 [0] | (-.743,.330) | .450 | 334,659 |
| | 5 Units of BMI | 5 Units of alcohol per week | -.098 [0] | (-.383,.187) | -.342 [0] | (-.809,.124) | .060 [0] | (-.021,.142) | .148 | 251,837 |
| | 5 Units of BMI | SD Years of education | -.225 [0] | (-.753,.302) | -.786 [0] | (-1.789,.216) | .117 [0] | (-.065,.300) | .209 | 332,707 |
| | SD Lifetime smoking index | 5 Units of alcohol per week | .094 [0] | (-.474,.663) | -.005 [0] | (-.097,.087) | .013 [0] | (-.115,.141) | .842 | 251,726 |
| | SD Lifetime smoking index | SD Years of education | .572 [0] | (-.265,1.408) | -.038 [0] | (-.195,.120) | -.160 [0] | (-.471,.151) | .312 | 332,689 |
| | 5 Units of alcohol per week | SD Years of education | .029 [0] | (-.329,.387) | -.134 [0] | (-.579,.310) | -.007 [0] | (-.125,.111) | .903 | 250,436 |

*Risk difference for interaction coefficient from linear regression, adjusted for sex, age, centre, PCs, exposure 1 and exposure 2 (Multivariate regression) or Meta-analysed estimate of interaction coefficient from two two-stage least squares estimates from split-sample analyses, see manuscript for model information (MR)
**Alcohol excluded from multimorbidity definition when it is part of the interaction

#### Table 5: IVW, MR-Egger, Simple Modal and Unweighted Median Estimates of Associations of Exposures with Multimorbidity Status (2+ conditions)

| Exposure | Outcome | Split | IVW | | | MR-Egger Slope | | MR-Egger Constant | | Simple Modal | | Unweighted Median | |
|---|---|---|---|---|---|---|---|---|---|---|---|---|---|
| | | | Beta (3.d.p) [SE] (3.d.p) | p-value (3.d.p) | Heterogeneity p-value (3.d.p.) | Beta (3.d.p) [SE] (3.d.p) | p-value (3.d.p) | Constant (3.d.p) [SE] (3.d.p) | p-value (3.d.p) | Beta (3.d.p) [SE] (3.d.p) | p-value (3.d.p) | Beta (3.d.p) [SE] (3.d.p) | p-value (3.d.p) |
| SD Years of education | 2+ conditions | 1 | -.062[.016] | <0.001 | <0.001 | .077[.114] | .500 | -.003[.002] | .218 | -.035[.043] | .412 | -.051[.018] | .005 |
| | | 2 | -.084[.014] | <0.001 | .025 | -.140[.079] | .077 | .001[.002] | .469 | -.051[.038] | .188 | -.063[.017] | <0.001 |
| | | Combined | -.075[.011] | <0.001 | NA | -.070[.065] | .285 | 0.000[.001] | .907 | NA | NA | NA | NA |
| 5 Units of BMI | 2+ conditions | 1 | .004[0.000] | <0.001 | <0.001 | .002[.001] | .037 | .001[.001] | .037 | .004[.001] | .001 | .004[0.000] | <0.001 |
| | | 2 | .003[0.000] | <0.001 | <0.001 | .001[.001] | .178 | .001[.001] | .072 | .003[.001] | .026 | .003[0.000] | <0.001 |
| | | Combined | .003[0.000] | <0.001 | NA | .002[.001] | .014 | .001[0.000] | .006 | NA | NA | NA | NA |
| SD Lifetime smoking index | 2+ conditions | 1 | .169[.049] | <0.001 | .006 | -.133[.147] | .368 | .003[.002] | .031 | .206[.110] | .060 | .167[.056] | .003 |
| | | 2 | .134[.046] | .003 | .046 | -.100[.171] | .559 | .003[.002] | .158 | .327[.125] | .009 | .158[.062] | .011 |
| | | Combined | .150[.033] | <0.001 | NA | -.119[.112] | .287 | .003[.001] | .011 | NA | NA | NA | NA |
| 5 Units of alcohol per week | 2+ conditions (excluding alcohol) | 1 | 0.000[0.000] | .547 | <0.001 | 0.000[.001] | .575 | -.001[.002] | .785 | 0.000[.001] | .884 | 0.000[0.000] | .965 |
| | | 2 | 0.000[0.000] | .538 | .266 | 0.000[0.000] | .475 | -.002[.001] | .130 | -.001[.001] | .364 | 0.000[0.000] | .272 |
| | | Combined | 0.000[0.000] | .804 | NA | 0.000[0.000] | .378 | -.002[.001] | .142 | NA | NA | NA | NA |
| Analyses adjusted for age, sex and PCs<br>Meta-analysis performed for IVW and MR-Egger only | | | | | | | | | | | | | |

### Figures

*Figure 1:* Mediation of the educational inequality in multimorbidity (IE=Indirect Effect)

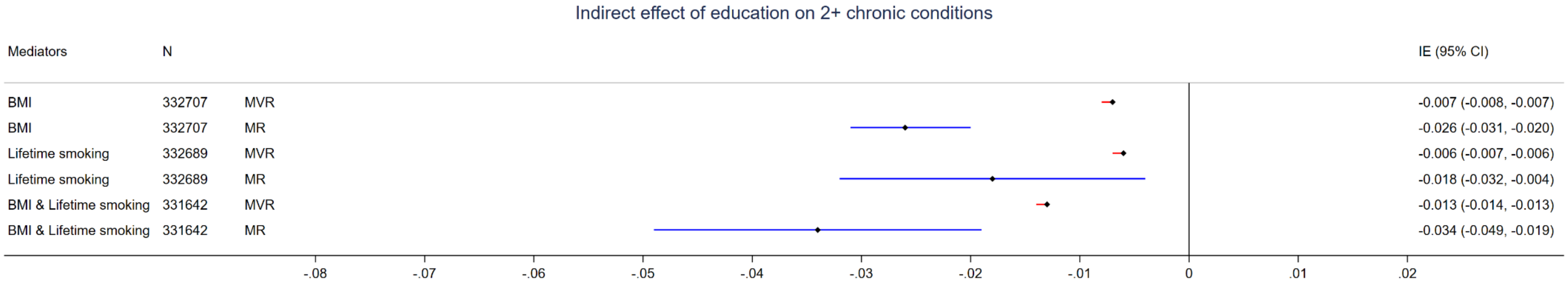
